# COVID-19 Pandemic in University Hospital: Impact on Medical Training of Medical Interns

**DOI:** 10.1101/2020.10.01.20204255

**Authors:** WeiHonn Lim, Li Ying Teoh, Kanesh Kumaran A/L Seevalingam, Shanggar Kuppusamy

## Abstract

**Introduction:** Coronavirus 2019 (COVID-19) has strike all nations hard since the end of year 2019, Malaysia unable to escape the fate as well. Healthcare system, financial growth, industrial development and educational programme are stunted. Inevitably, professional training and education are affected which include the medical training of medical interns.

**Methods:** This is a cross-sectional, pilot study to determine the impact of the pandemic on University Malaya Medical Centre (UMMC) medical interns. A survey which comprises 37-items was used. Data are analysed by Ordinal Logistic Regression Analysis.

**Results:** Medical interns feel that they lack clinical skills (p = 0.005) and need more exposure in surgical operations (p =0.029). Some are satisfied with the introduction of triage (p = 0.024), online teaching (p = 0.005) and bedside teaching (p=0.023). Most of them think they are fit and ready to handle the pandemic (p = 0.012 and 0.025 respectively) except first year medical interns (p = 0.029). Some feel like their time are wasted (p <0.05) as they are involved in many non-clinical activities (p = 0.003).

**Conclusion:** In summary, COVID-19 has a great impact on medical training amongst medical interns. Alternative measures should be taken to minimize the interruption in training of our future leaders in medical field.

## Introduction/Background

COVID-19 mercilessly invades the globe with its virus and causes a havoc globally. [1] In order to face this pandemic crisis, healthcare field has suffered the most with its limited resources, burnt-out healthcare workers (HCW) and sudden raise in workloads.[2, 3] Malaysia unavoidably suffered similar fate since March 2020 and received a massive hit from this pandemic. In response to this pandemic crisis, Malaysia has implemented Movement Control Order (MCO) from 18^th^ March 2020 in several stages, and currently we are in the Recovery Period of MCO.[4] To curb the spreading of COVID-19, several measures are implemented to contain the disease by alternating the current medical training from undergraduate to medical interns: rescheduling clinic appointment and elective operations to reduce unaffected populations from contracting COVID-19[5-7]; reshuffling HCW to overcome shortage of staffs especially in Emergency Department; suspension of bedside clinical teaching to avoid transmission within HCW; cancellation of workshop and conference to avoid mass gathering.[8, 9] Inevitably, medical training for medical interns are affected. [10, 11]

The medical interns, also known as the House Officers in Malaysia, comprise of medical fresh graduates who must complete their two-year medical training. During this period of training, they rotate to six compulsory clinical postings namely Internal Medicine, Surgery, Obstetrics and Gynaecology, Orthopaedic and Paediatric departments. Addition to that is a choice of a posting in either Psychiatry, Emergency Medicine or Anaesthesiology department. They are required to complete at least four months rotation in each department with a certain expectation in their clinical exposure to be fulfilled under full supervision by their specialists and consultants. [12]This two-year medical training is tough and medical interns have to withstand a great level great level of work stress while trying to gain more clinical experiences during the posting in each department.[13, 14]

As a teaching university hospital in Malaysia, University Malaya Medical Centre (UMMC) has been gazetted as one of the designated COVID centres. To fulfil its role, many changes have been adapted to accommodate the surge of COVID cases: from rescheduling clinic appointments and elective operations to deployment of more HCW to Emergency Department as “frontliners” and gazetting more designated COVID wards to manage infected cases. [15] Furthermore, due to shortage of medical equipment such as personal protective equipment (PPE) faced by UMMC initially, limitation to numbers of healthcare workers to have direct contact with patients were issued. Indirectly, medical interns who are supposed to gain knowledge and experiences in managing patients are greatly affected.[10] Additionally, they have to work under pressures, heighten their vigilance during their works and worry about getting inflicted with diseases. All these invisible hurdles not only affected their mental states but their performances and their ability to learn. Nevertheless, the degree of impact from this pandemic on the medical interns needs to be addressed to ensure a continuous and effective clinical training.

## Methodology

A survey form is created and drafted in English language which composed of two components: demographic status (gender, ethnics, age, number of posting and qualification attained) and the questionnaire itself which comprised of five domains: physical health, emotion/ psychological coping, social/ relationship handling, medical training progression and preparedness in handling a pandemic situation. The questionnaire uses a five-response Likert Scale: ranging from 1 to 5 which represent totally disagree, disagree, neutral, agree and strongly agree respectively.

The questionnaire is uploaded to an online survey form - Google Form and a link is generated. The invitation for participation in the study is sent by email and electronic devices via WhatsApp, with the link to assess the questionnaire. Electronic Participant Information Sheets (PIS) are provided in the first page of the survey form. All participations are self-voluntary whereby they can choose not to participate if they have disagreed to include their responses in this study result. The questionnaire is self-administered by the participants in anonymity. Participation is opened from April 2020 to June 2020.

Subsequently, all data are downloaded into Excel form and transcript into SPSS Version 16.0. Demographic data are summarised by descriptive analysis. Association between demographic factors and each item in the questionnaire is analysed by using Ordinal Logistic Regression Analysis. Subsequently, data are analysed using Generalised Linear Models where demographic data were entered as the factor and all items in the questionnaire were keyed in as dependent variables. For each factor, groups with majority numbers are made as comparators to avoid potential bias: Female, Malay ethnic and local graduates are listed as comparators to others in their respective groups.

## Results

There are total of 236 responses recorded in our Google Form. Out of these numbers, 232 participants consented to join which contributes 98.3% of response rate while only four of them declined to participate. Of 232 participants who agreed to participate in this survey, majority are females (62.1%) and of Malay ethnics (45.3%). Most of them attained their medical degrees from local university (60.8%) and had completed at least 3 postings (56.9%).

**Table 1:** Basic Demographic Data of Medical Interns

| *Variables* | *Male*<br>(*n* = 88) | *Female*<br>(*n* = 144) | *Total*<br>(*n* = 232) |
| --- | --- | --- | --- |
| **Age, years (SD)** | **27 (1.38)** | **27 (1.74)** | **27 (1.6)** |
| **Ethnics, *n* (%)** |  |  |  |
| Malay | 33 (14.2) | 72 (31.0) | 105 (45.3) |
| Chinese | 35 (15.1) | 43 (18.5) | 78 (33.6) |
| Indian | 17 (7.3) | 24 (10.3) | 41 (17.7) |
| Others | 3 (1.3) | 5 (2.2) | 8 (3.4) |
| **Qualification Attained, *n* (%)** |  |  |  |
| Local University | 56 (24.1) | 85 (36.6) | 141 (60.8) |
| Overseas University | 18 (7.8) | 44 (19.0) | 62 (26.7) |
| Twinning Programmes | 14 (6.0) | 15 (6.5) | 29 (12.5) |
| **Number of Posting, *n* (%)** |  |  |  |
| 1 | 13 (5.6) | 14 (6.0) | 27 (11.6) |
| 2 | 14 (6.0) | 25 (10.8) | 39 (16.8) |
| 3 | 24 (10.3) | 42 (18.1) | 66 (28.4) |
| 4 | 20 (8.6) | 36 (15.5) | 56 (24.1) |
| 5 | 6 (2.6) | 11 (4.7) | 17 (7.3) |
| 6 | 11 (4.7) | 16 (6.9) | 27 (11.6) |

**Table 2:** Association between Demographic Factors with Medical Training Progress

| Questions | Gender |  | Ethnic |  | Location of Qualification |  | Number of Posting |  |
| --- | --- | --- | --- | --- | --- | --- | --- | --- |
|  | Estimates<br>(OR) | p-value<br>(Confidence<br>Interval) | Estimates<br>(OR) | p-value<br>(Confidence<br>Interval) | Estimates<br>(OR) | p-value<br>(Confidence<br>Interval) | Estimates<br>(OR) | p-value<br>(Confidence<br>Interval) |
| I feel that I am lacking experiences in hands-on procedures. | -0.843<br>(0.431) | 0.001<br>(0.262-0.707) | E1 = -1.703<br>(0.182) | 0.100<br>(0.050-0.660) | G1 = 0.473<br>(1.605) | 0.197<br>(0.782-3.294) | P1 = -0.276<br>(0.759) | 0.586<br>(0.281-2.049) |
|  |  |  | E2 = -0.743<br>(0.476) | 0.031<br>(0.242-0.936) | G2 = 0.801<br>(2.228) | 0.005<br>(1.273-3.901) | P2 = -0.608<br>(0.544) | 0.185<br>(0.221-1.339) |
|  |  |  | E3 = -0.970<br>(0.907) | 0.720<br>(0.534-1.542) |  |  | P3 = -0.861<br>(0.423) | 0.040<br>(0.186-0.961) |
|  |  |  |  |  |  |  | P4 = 0.202<br>(1.224) | 0.635<br>(0.532-2.817) |
|  |  |  |  |  |  |  | P5 = 0.430<br>(1.538) | 0.484<br>(0.460-5.138) |
| I wish we had more surgical emergency operations. | -0.567<br>(0.567) | 0.024<br>(0.347-0.928) | E1 = -1.335<br>(0.263) | 0.055<br>(0.067-1.027) | G1 = 0.253<br>(1.288) | 0.504<br>(0.612-2.709) | P1 = -1.118<br>(0.327) | 0.024<br>(0.124-0.861) |
|  |  |  | E2 = -0.096 | 0.776 | G2 = 0.626 | 0.029 | P2 = -1.176 | 0.009 |
|  |  |  | (0.909) | (0.470<br>-1.758) | (1.871) | (1.064-3.288) | (0.308) | (0.127-0.747) |
|  |  |  | E3 = -0.216<br>(0.806) | 0.432<br>(0.471-1.380) |  |  | P3 = -0.768<br>(0.464) | 0.064<br>(0.206-1.047) |
|  |  |  |  |  |  |  | P4 = -0.160<br>(0.852) | 0.710<br>(0.367-1.977) |
|  |  |  |  |  |  |  | P5 = -0.815<br>(0.443) | 0.155<br>(0.144-1.361) |
| I am satisfied with the current<br>online teaching for HOs<br>(department/division level). | 0.059<br>(1.061) | 0.812<br>(0.653-1.722) | E1 = 0.971<br>(2.639) | 0.180 (0.640-10.888) | G1 = -<br>0.222<br>(0.801) | 0.553<br>(0.386-1.665) | P1 = 0.634<br>(1.884) | 0.214<br>(0.694-5.116) |
|  |  |  | E2 = -0.278<br>(0.758) | 0.409<br>(0.392-1.465) | G2 = -<br>0.142<br>(0.868) | 0.610<br>(0.502-1.498) | P2 = 0.543<br>(1.721) | 0.240<br>(0.695-4.257) |
|  |  |  | E3 = -0.513<br>(0.599) | 0.065<br>(0.347-1.033) |  |  | P3 = 1.227<br>(3.411) | 0.005<br>(1.455-7.996) |
|  |  |  |  |  |  |  | P4 = 0.151<br>(1.163) | 0.728<br>(0.495-2.732) |
|  |  |  |  |  |  |  | P5 = 0.395 | 0.505 |
|  |  |  |  |  |  |  | (1.485) | (0.464-4.747) |
| I do not receive adequate bedside teaching from my MOs/Lecturers/Consultants | -0.303<br>(0.739) | 0.213<br>(0.459-1.190) | E1 = -0.883<br>(0.414) | 0.212<br>(0.103-1.654) | G1 = 0.358<br>(1.430) | 0.328<br>(0.698-2.932) | P1 = 0.031<br>(1.032) | 0.950<br>(0.390-2.729) |
|  |  |  | E2 = -0.307<br>(0.736) | 0.372<br>(0.375-1.443) | G2 = 0.490<br>(1.632) | 0.082<br>(0.940-2.835) | P2 = -0.286<br>(0.751) | 0.535<br>(0.304-1.854) |
|  |  |  | E3 = 0.235<br>(1.265) | 0.376<br>(0.752-2.126) |  |  | P3 = -0.963<br>(0.382) | 0.023<br>(0.166-0.875) |
|  |  |  |  |  |  |  | P4 = -0.107<br>(0.898) | 0.803<br>(0.387-2.084) |
|  |  |  |  |  |  |  | P5 = -0.523<br>(0.593) | 0.360<br>(0.194-1.814) |
| I learn about the triage and management of patients during a pandemic. | 0.422<br>(1.524) | 0.095<br>(0.930-2.499) | E1 = 0.256<br>(1.291) | 0.688<br>(0.371-4.493) | G1 = 0.146<br>(1.157) | 0.700<br>(0.550-2.433) | P1 = -0.935<br>(0.393) | 0.072<br>(0.142-1.087) |
|  |  |  | E2 = 0.777<br>(2.175) | 0.024<br>(1.108-4.270) | G2 = -<br>0.727<br>(0.484) | 0.010<br>(0.278-0.842) | P2 = -0.675<br>(0.509) | 0.159<br>(0.199-1.301) |
|  |  |  | E3 = 0.152<br>(1.165) | 0.579<br>(0.679-1.997) |  |  | P3 = 0.026<br>(1.026) | 0.953<br>(0.431-2.445) |
|  |  |  |  |  |  |  | P4 = -1.064 | 0.020 |
|  |  |  |  |  |  |  | (0.345) | (0.141-0.844) |
|  |  |  |  |  |  |  | P5 = 0.539<br>(1.714) | 0.375<br>(0.521-5.634) |
| I have been exposed to the<br>importance of audit/research<br>during this pandemic. | 0.023<br>(1.023) | 0.928<br>(0.624-1.676) | E1 = 0.139<br>(1.149) | 0.830<br>(0.322-4.096) | G1 = 0.129<br>(1.137) | 0.727<br>(0.552-2.341) | P1 = -0.089<br>(0.914) | 0.862<br>(0.334-2.506) |
|  |  |  | E2 = 0.402<br>(1.495) | 0.249<br>(0.754-2.964) | G2 = 0.080<br>(1.083) | 0.776<br>(0.624-1.881) | P2 = -0.294<br>(0.745) | 0.519<br>(0.305-1.820) |
|  |  |  | E3 = -0.176<br>(0.839) | 0.520<br>(0.490-1.434) |  |  | P3 = 0.742<br>(2.101) | 0.082<br>(0.909-4.856) |
|  |  |  |  |  |  |  | P4 = -0.038<br>(0.963) | 0.931<br>(0.412-2.251) |
|  |  |  |  |  |  |  | P5 = 0.877<br>(2.402) | 0.149<br>(0.731-7.896) |

\*E1 = other ethnics; E2 = Indian; E3 = Chinese; G1 = twining program; G2 = overseas; P1= 1<sup>st</sup> posting; P2 = 2<sup>nd</sup> posting; P3= 3<sup>rd</sup> posting; P4 = 4<sup>th</sup> posting; P5 = 5<sup>th</sup> posting; HO =house officer; MO = medical officer

**Table 3:**
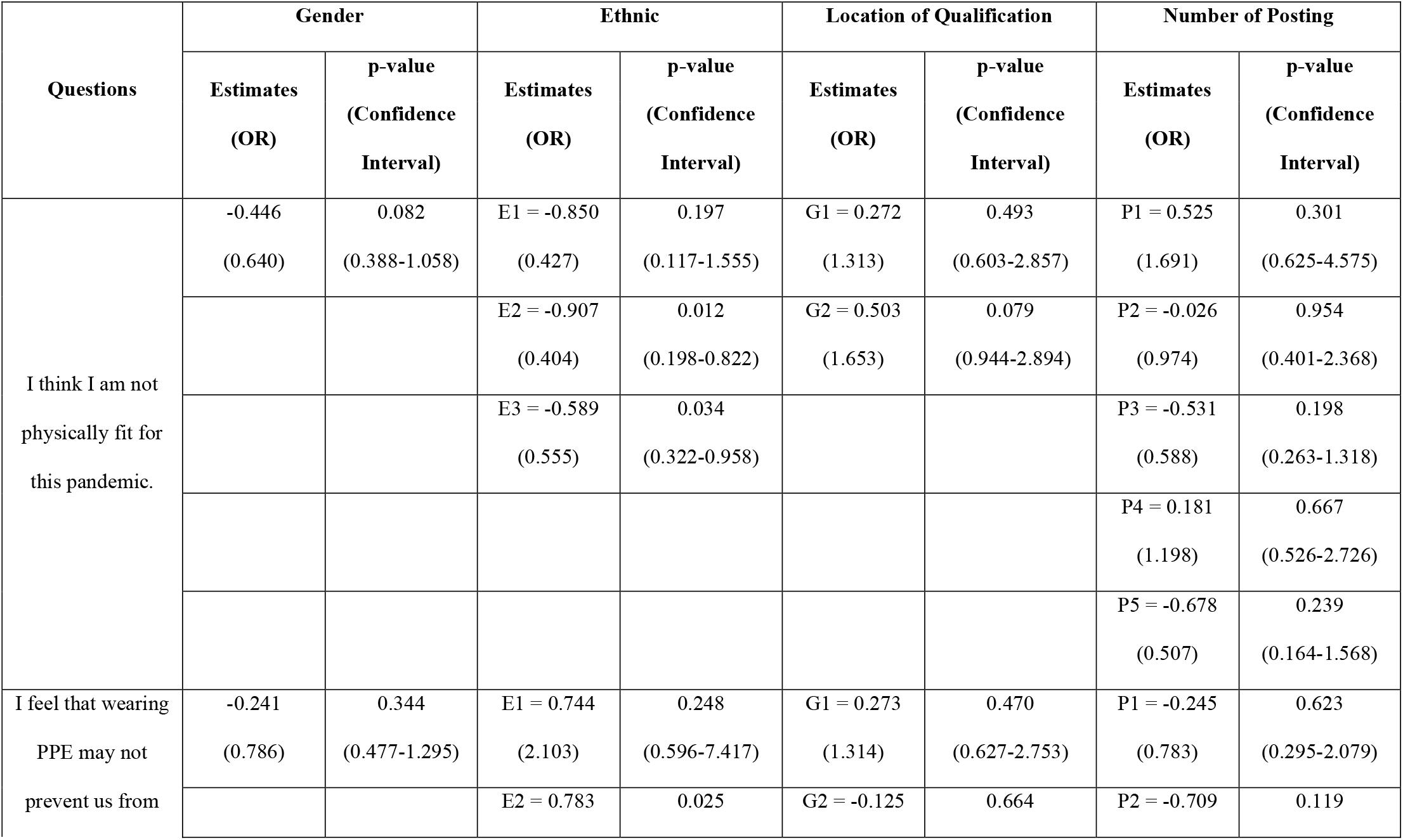
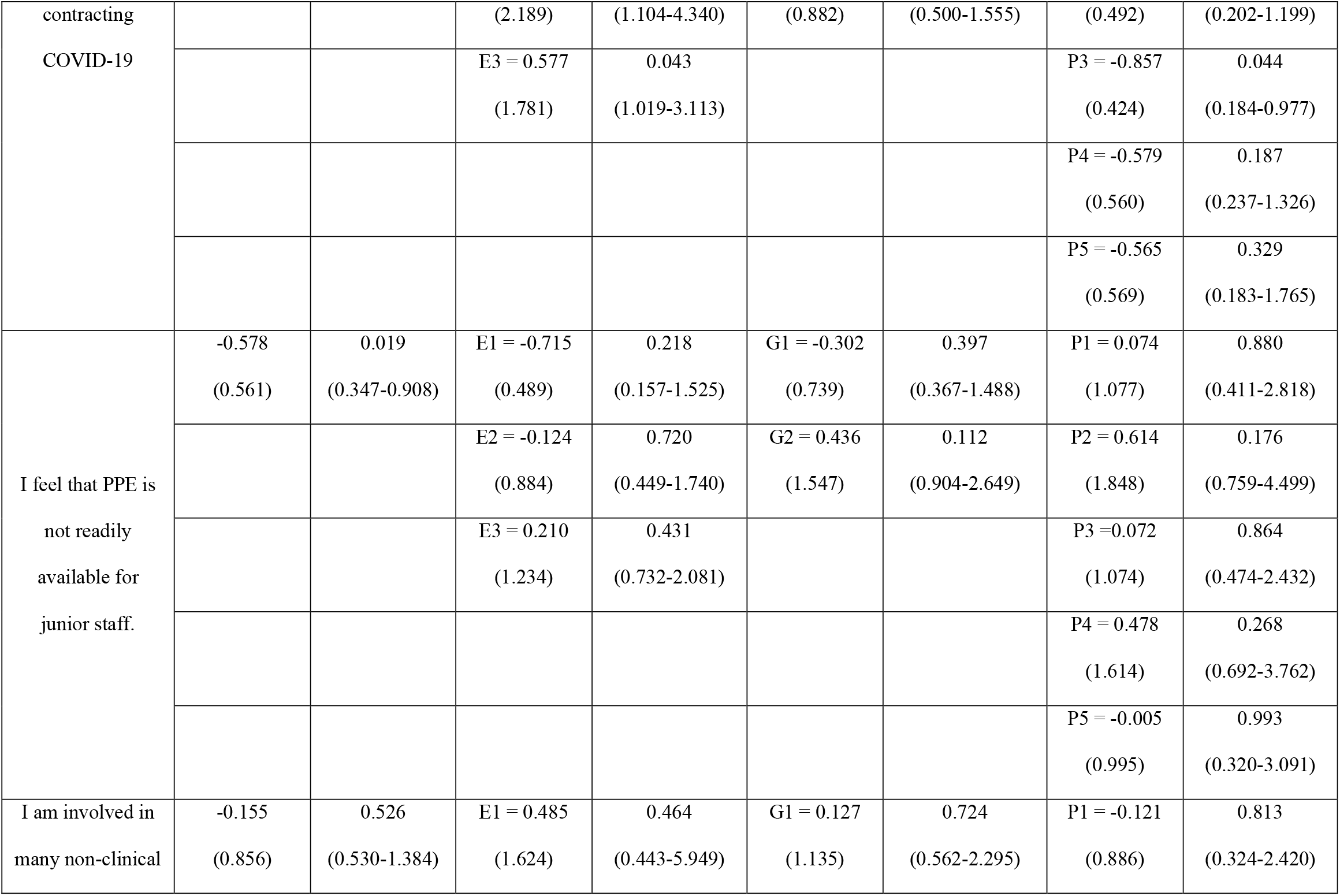
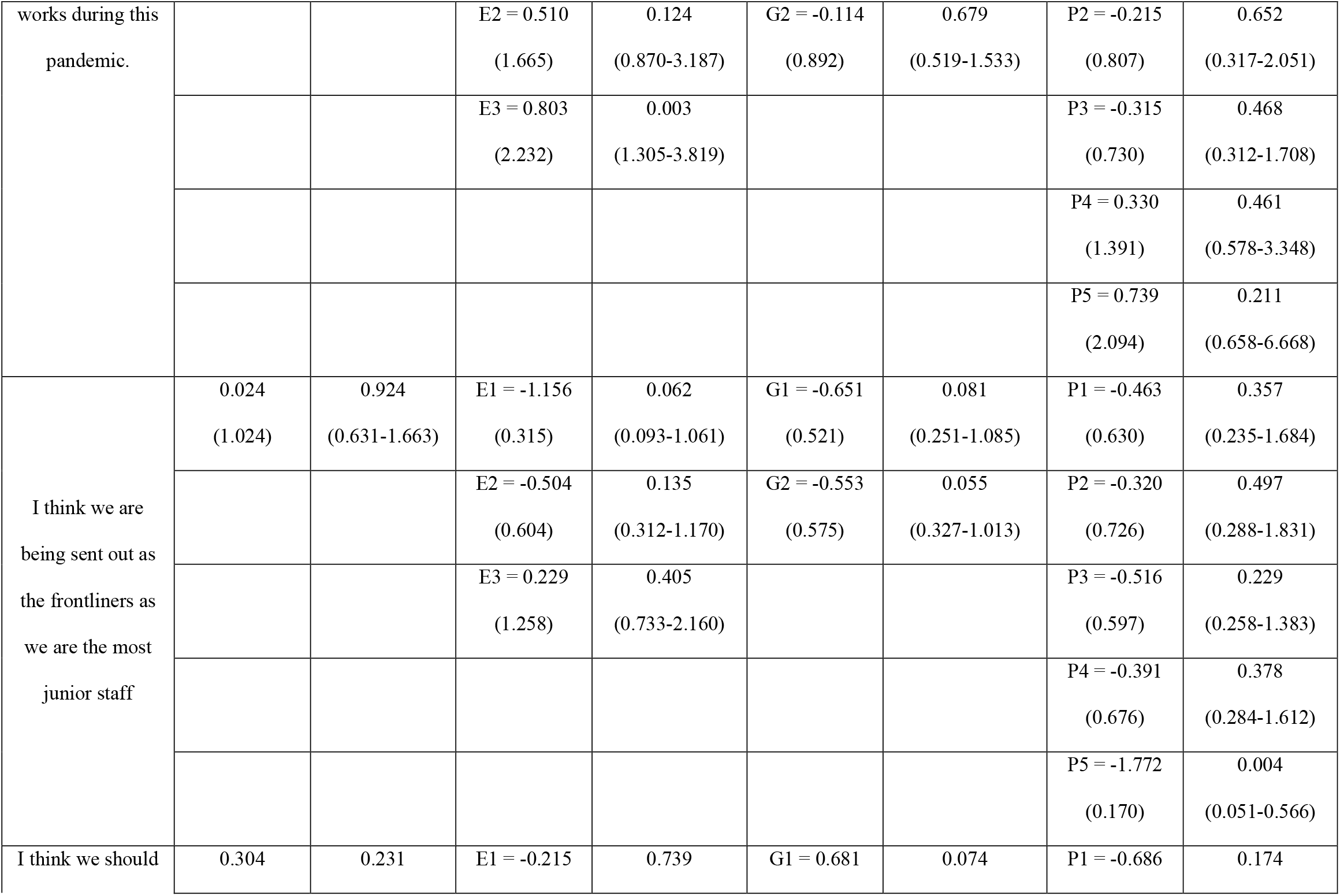
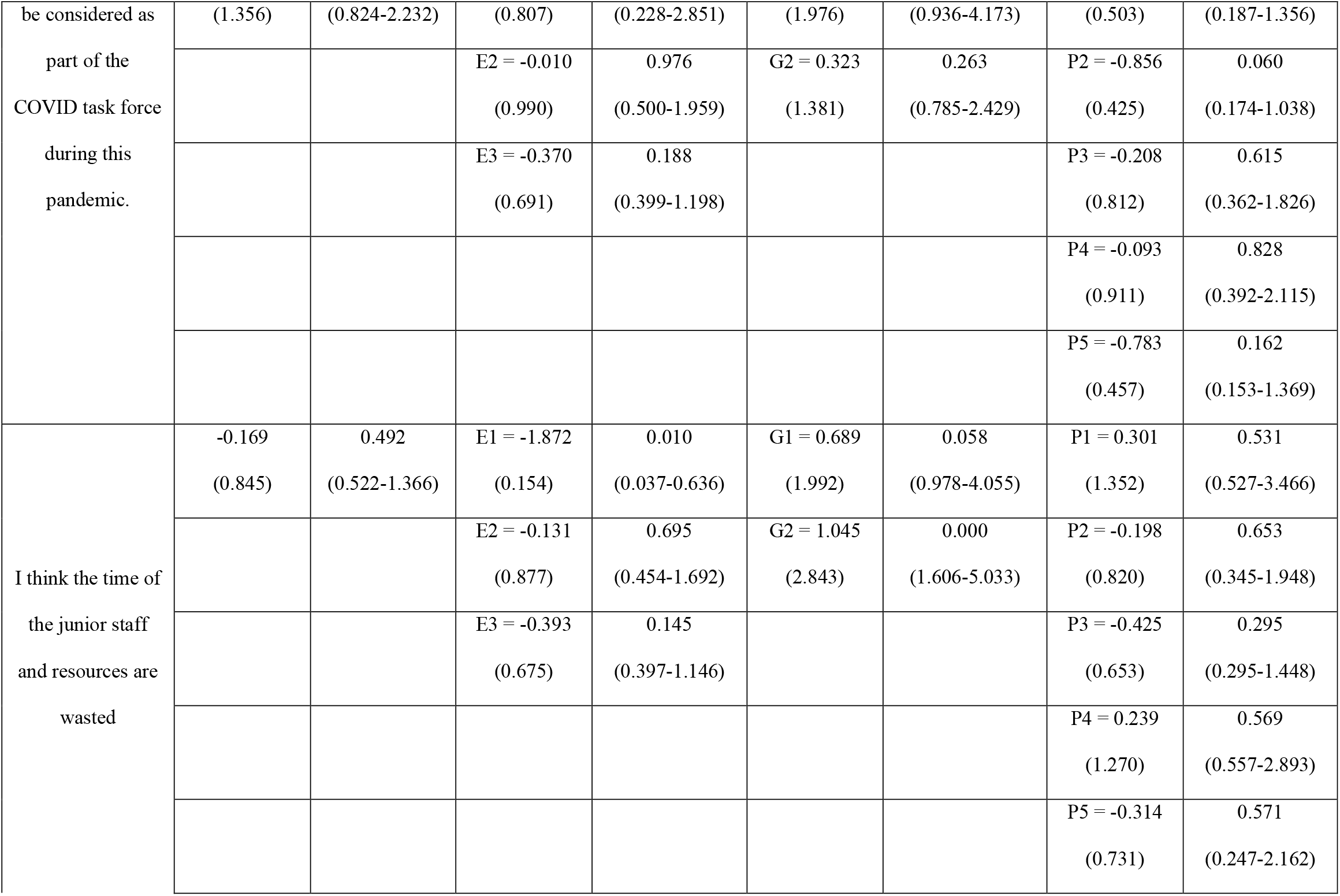
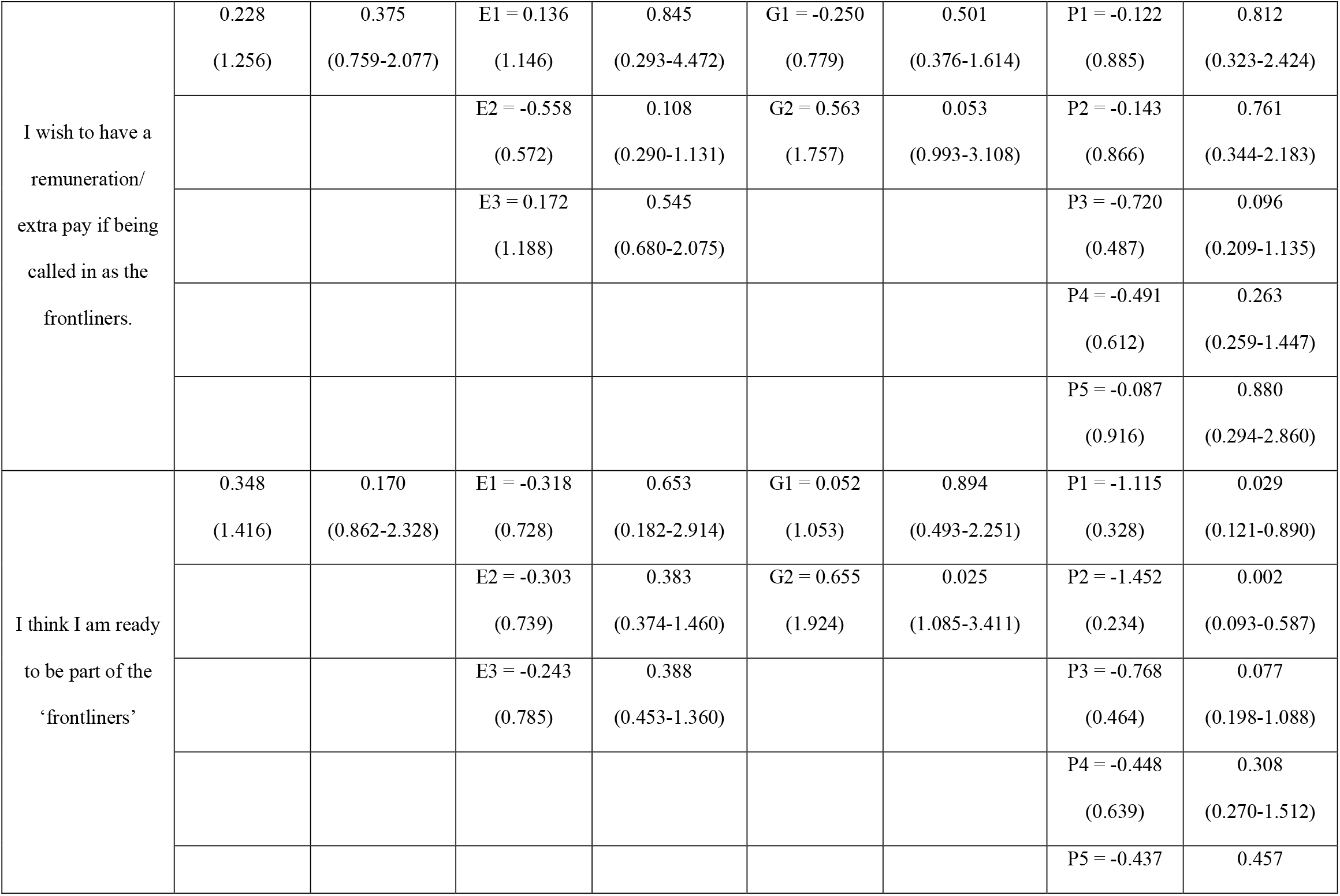
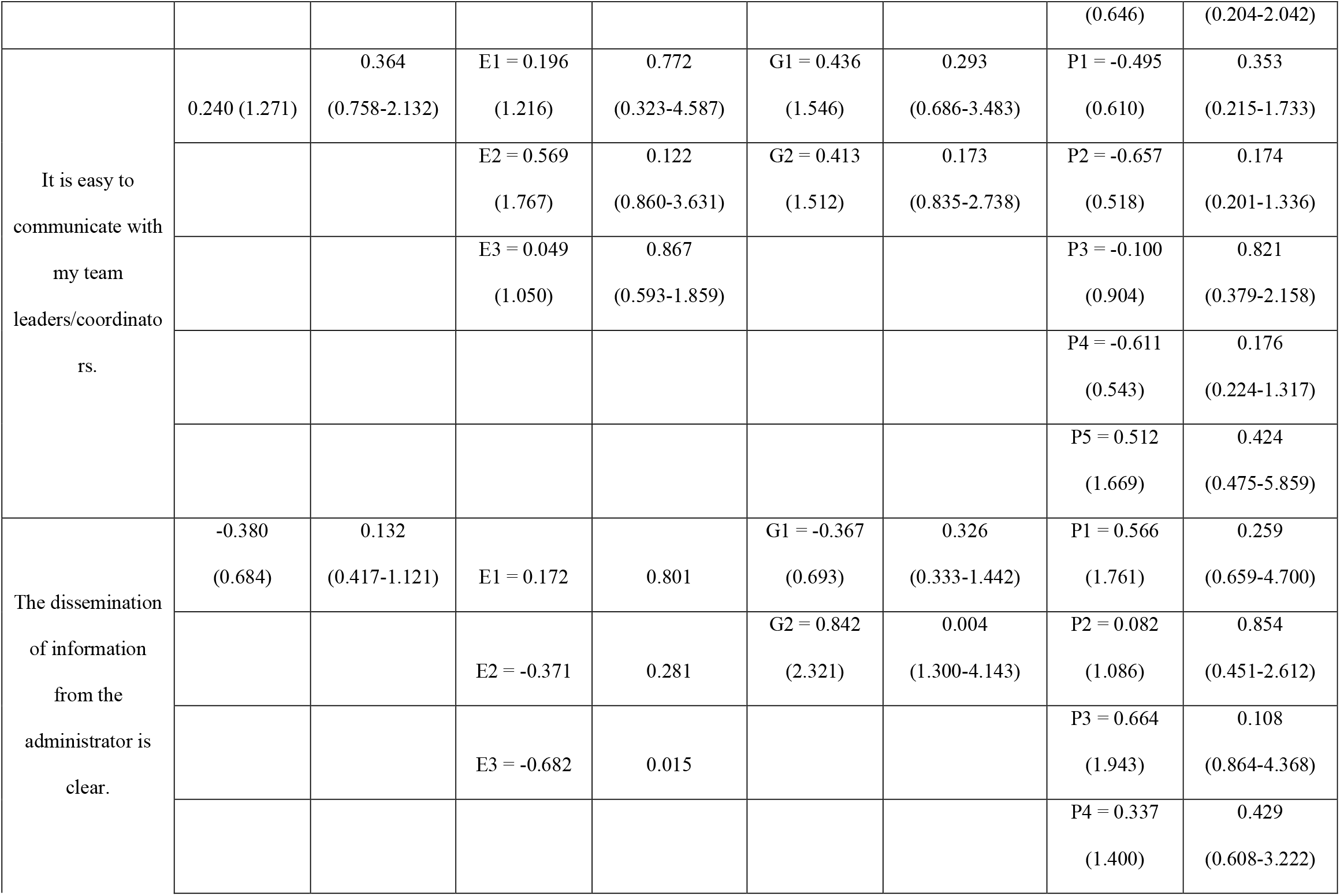
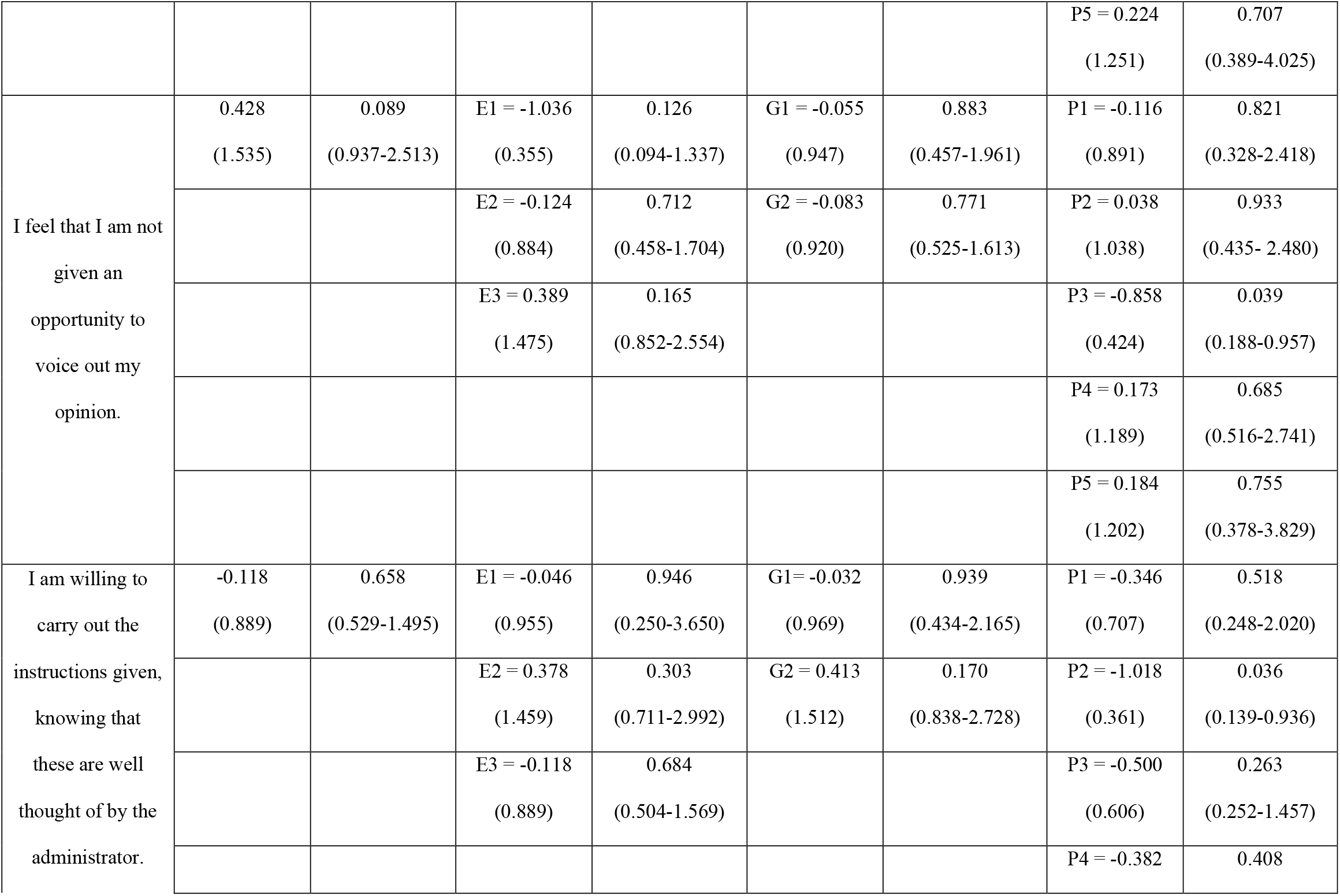
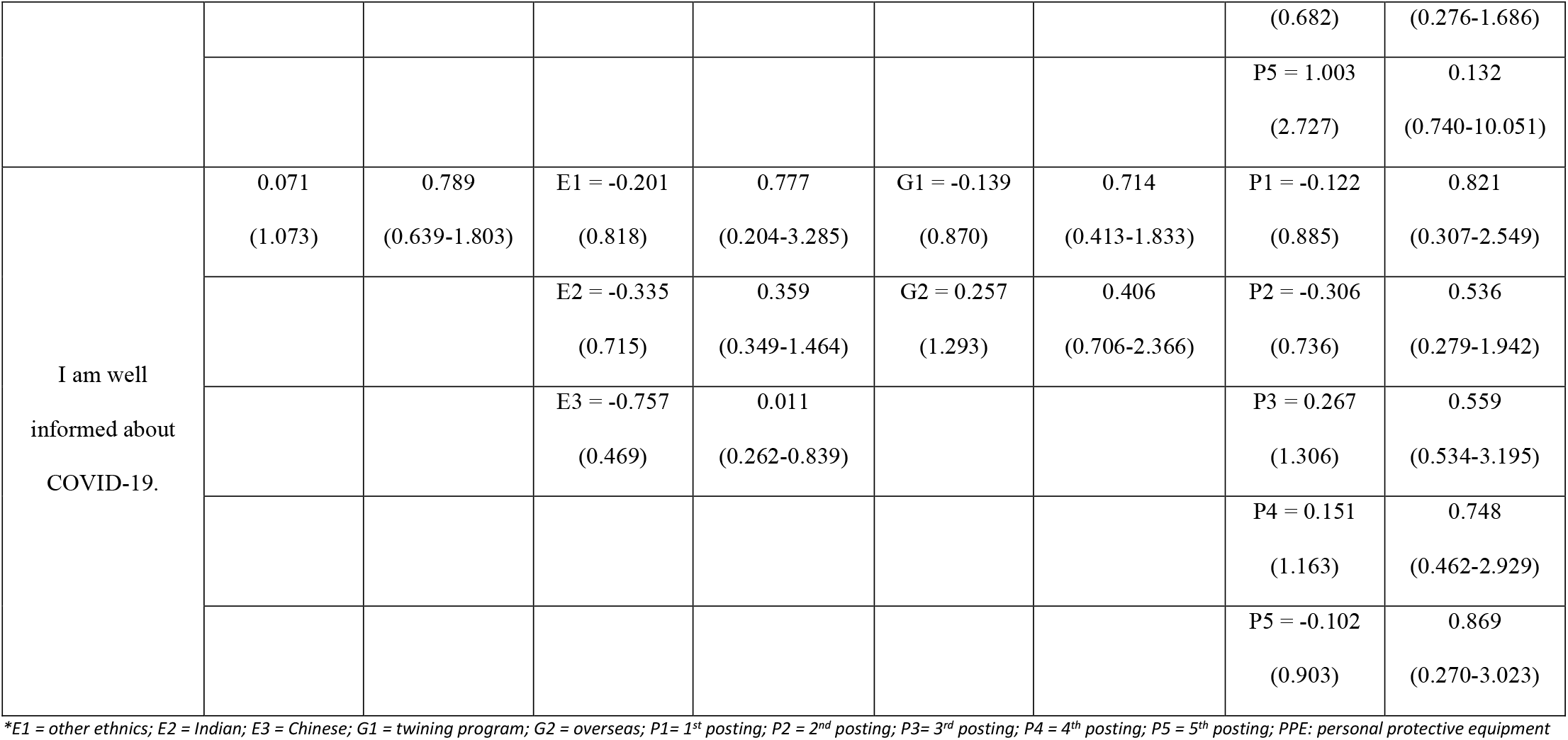
Association between Demographic with Preparedness in Handling A Pandemic

### Medical Training Progress

From the survey, majority of the medical interns who graduated from overseas think that they are lack of skill [p = 0.005, OR = 2.228 (95% CI 1.273 - 3.901)] and need more surgical procedures [p = 0.029, OR = 1.871 (95% CI 1.064 - 3.288)] Surprisingly, male medical interns do not think they are lack of experience in hands-on skills [p = 0.001, OR = 0.431 (95% CI 0.262 - 0.707)] and do no need more surgery operations [p= 0.024, OR = 0.567 (95% CI 0.347 - 0.928)]; Indians also think do not think they lack of skill [p = 0.031, OR = 0.476 (95% CI 0.242 - 0.936)] Based on number of posting, third posting do not think they lack of experience too[p = 0.040, OR = 0.423 (95% CI 0.186 - 0.9610)]. And junior medical interns (first and second posting) do not feel they need more surgical operations [(P1) p = 0.024, OR = 0.327 (95% CI 0.124 - 0.861)/ (P2) p = 0.009, OR = 0.308 (95% CI 0.127 - 0.747)]

During this COVID period, many Indian medical interns have learnt the importance of triage and management of patients [p = 0.024, OR = 2.175 (95% CI 1.108 - 4.270)] but not for those graduated from overseas [p = 0.010, OR = 0.484 (95% CI 0.278 - 0.842)] and fourth posting medical interns [p = 0.020, OR = 0.345 (95% CI 0.141 - 0.844)]

Many are satisfied with online teaching, especially third posting [p = 0.005, OR = 3.411 (95% CI 1.455 - 7.996)] and they think there is adequate of bedside teaching [p = 0.023, OR = 0.382 (95% CI 0.166 - 0.875)]

### Preparedness in Handling Pandemic Situation

Most guys think PPE are readily available for medical interns [p = 0.019, OR = 0.561 (95% CI 0.347 - 0.908)] On the other hands, medical interns from third posting feel that by wearing PPE it will prevent them from infection [p = 0.044, OR = 0.424 (95% CI 0.184 - 0.977)] but Indian medical students do not think so [p = 0.025, OR = 2.189 (95% CI 1.104 - 4.340)]

Medical interns who graduated from overseas think the time of the junior staffs and resources are wasted [p = 0.000, OR = 2.843 (95%CI 1.606 - 5.033)] while some of think they are involved in many non-clinical stuff during this pandemic, especially Chinese [p = 0.003, OR = 2.232 (95% CI 1.305 - 3.819)]

From the survey, we noticed most of the medical interns feel they are physically fit for this pandemic, especially Chinese and Indians [(Chinese) p = 0.012, OR = 0.404 (95% CI 0.198 - 0.822)/ (Indian) p = 0.034, OR = 0.555 (95% CI 0.322 - 0.958)] Most of the medical interns who graduated from overseas think they are ready to be frontliners [p = 0.025, OR = 1.924 (95% CI 1.085 - 3.411)] but junior medical interns from first and second posting feel they are not ready to be frontliners yet [(P1) p = 0.029, OR = 0.328 (95% CI 0.121 - 0.890)/ (P2) p=0.002, OR = 0.234 (95% CI 0.093 - 0.587)]

For the dissemination of information during this pandemic crisis, majority of Chinese medical interns think the distribution is unclear [p = 0.015, OR = 0.506 (95% CI 0.292 - 0.875)] and they are not well-informed [p = 0.011, OR = 0.469 (95% CI 0.262 - 0.839)]. However, medical interns who graduated from overseas do not think so [p = 0.004, OR = 2.321 (95% CI 1.300 - 4.143)] Most of the medical interns think they are not given an opportunity to voice out their opinion except third posting [p = 0.039, OR = 0.424 (95% CI 0.188 - 0.957)]. Unfortunately, many of them do not agree with the statement of “I am willing to carry out the instructions given, knowing that these are well thought of by the administrator” especially second posting [P = 0.036, OR = 0.361 (95% CI 0.139 - 0.936)] Senior medical interns especially fifth posting do not think they are being sent out as the frontlines as they are the most junior staff [p = 0.004, OR = 0.170 (95% CI 0.051 - 0.566)]

## Discussion

House Officers (HO) are medical interns who just graduated from medical colleges or universities. In Malaysia, they will undergo two years of internship and be assigned to six different postings with duration of 4 months for each posting. The purpose of these two-year training is to equip them with sufficient clinical skills and experiences to manage patients. Without completion of training, they are not allowed to practice medicine. Being part of the training is stressful as they must go through different fields and learn to manage patients in respective fields within limited time. Furthermore, they have pressures from superiors, expectation from patients and requirements to be fulfilled by performing numbers of basic procedures within that short period of time. [13, 14]

COVID-19 pandemic has affected all HCW greatly in terms of routine ward rounds, elective operations and clinic arrangement, and disrupted the healthcare system. Additionally, they are suffering from immense stresses as they are worried of contracting the virus while treating the patients.[16-18] In addition, educational training and medical teaching session have ceased until further notice.[9, 11, 19] Hands-on workshops and conferences are prohibited during this pandemic period. They had inadequate cases and procedures to observe; facing lack of experience and hands-on clinical skills; being re-allocated depending on capacity and workload of the hospital. [5, 10]Facing tremendous uncertainties during this pandemic crisis, medical interns as the junior most doctors face far more pressures than other HCW.

### Medical Training Progress

During COVID pandemic, many surgeries include elective operations have been rescheduled to accommodate COVID patients. Thus, less patients will be admitted for surgeries which lead to less procedures and operations. [6, 10] Many medical interns who are graduated from overseas feel that they lack experience and wish for more hands-on procedures. They realised the importance of training for their future undertakings as well as being a qualified doctor requires not only knowledge, but clinical skills too. Presence of such thoughts might attribute to their early exposure to modern medicine overseas which have broadened their views and increased the level of maturity. However, some medical interns do not think they lack experience and thus do not need more procedures or operations, mainly first year male medical interns. The reasons of junior medical interns do not feel they need more surgical procedures could be because they are still new to the medical field and do not understand the expectation of their trainings. Overwhelming workload during this COVID-19 season has dampened their interests.

As for educational teaching, third posting medical interns are satisfied with both online and bedside teaching conducted by seniors and lecturers. Most of them have adapted to the new routine ward round and thus, they find it convenient to join the teaching sessions especially in less hectic postings such as anaesthesiology and emergency departments. Thus, they can join the teaching sessions and attained work-learning balance during this period.

### Preparedness in Handling Pandemic Situation

Most of the medical interns think they are fit and ready to be part of the frontlines in dealing with COVID cases.[20] On contrary, junior medical interns, especially first posting, do not feel that they are ready yet. This could be explained as they are new to the system and lack of knowledge as well as clinical skills. They are still learning and adapting to current healthcare system. However, COVID-19 complicates the process and might demotivate the junior medical interns.

Some medical interns feel like their time and resources are wasted while others feel that they have involved in many non-clinical activities during this period. As a measure to protect medical interns and prevent local transmission between HCW, they are allocated in different clusters to handle different tasks which include ward rounds, rescheduling elective operations and clinic administration, data entry for census or research study and assisting Medical Officers in emergency department. Some might feel they should be in the “battlefields” with others in handling COVID cases. This shows most of the medical interns are enthusiastic to be part of the frontlines.

During a pandemic attack, dissemination of information is important as it provides the latest updates and possible measures to counter any problems faced. [21] Some medical interns feel that the information is unclear, and they are not being updated with the latest news. The reasons of having vague information of COVID-19 are contributed by the facts that the viral outbreak strikes rapidly across worldwide with the presence of different strains, making it difficult to completely understand its nature in a short period of time. Additionally, transmission of information from higher authorities to respective COVID centres does not keep up with the speed of viral spread. Up to date, there are still no vaccine or medicine to treat it. Most of the treatments focus on supportive management and this has shaken the confidence of HCW, including medical interns. In addition, they are not given chances to voice out their ideas and thoughts. Most of the time, ad-hoc plans and measures are channelled down by the higher authorities without clear explanation which might lead to the unsatisfaction among medical interns. Thus, some medical interns are not willing to abide the rules set by their superiors.

Fortunately, many feel that PPE is readily available even for junior staffs and they feel secure by wearing those PPE. Most of the countries suffer from limited resources due to the sudden demands worldwide. In dire situation, many HCW could not even have access to PPE while attending patients. [3, 22] Government and higher authorities have managed our resources well, ensuring equal distribution of the PPE so that all HCW are able to protect themselves during the treatment of the patients.

## Limitation

1. This is a pilot study where the questionnaire is not validated. The questionnaire is created based on problems encountered by medical interns during their daily routines.
2. As for the sample size, only 236 medical interns participate in this study with the initial calculated sample size of 378. This contributes to 63% of response rate[23][24] There are few problems identified which are some of the medical interns have finished their internship and had switched to other hospitals or awaiting placement; some of them are in the transition of switching departments; invalid email addresses; possibility of busy working schedules or being too preoccupied with their routine works.
3. In addition, this is a self-administered questionnaire in anonymity where repetition of entry is possible. Follow-up study could be helpful in assessing the progression of medical interns after COVID-19 pandemic subsides.

## Conclusion

COVID-19 pandemic has invaded many countries and paralysed most of the healthcare systems. During this challenging period, medical training for medical interns are affected.

Based on the results, medical interns are concerned of their competency as their medical trainings are affected during this pandemic crisis. For educational teaching, continuous medical educations (CME) should be continued in the form of online teaching. [9, 23] Various online applications could be used to provide effective teaching with good interaction between the educators and medical interns. By using similar methods, simulated procedures could be applied too. Bedside clinical teaching should be continued with some modification such as forming a smaller group of medical interns with adequate PPE and adherence to hand hygiene.[24] Introduction of non-clinical activities such as research and census are important but should not be an obstacle for the medical interns to learn and contribute to this pandemic.

As for the preparedness amongst medical interns in pandemic crisis, most of the medical interns think they are ready and should be included as part of the team.[20] They should be given a chance to join forces with other HCW in battling COVID-19. Encouragement and support should be provided to promote their growth in medicine without neglecting their safety and well-being. [25] With a proper guidance and distribution of information, we could turn this pandemic into an opportunity for them to learn and grow as they will be our future ray of hope if another pandemic strikes

In a nutshell, the impact of COVID-19 amongst medical interns is significant and should be addressed appropriately. Their overall well-being should be protected and preserved without jeopardise their trainings and preparation for pandemic crisis in the future.

## Data Availability

All data are available without restriction as presented in the manuscript.

## Abbreviations

COVID-19: Coronavirus Disease 2019
HCW: healthcare workers
UMMC: University Malaya Medical Centre
OR: Odd Ratio

## Acknowledgement

I sincerely would like to thank everyone who has contributed to this study. A heartful gratitude to all parties in helping to make this study a success:

1. House Officer Coordinators from different departments in UMMC
2. Human Resource Officers UMMC
3. Dr Retnagowri A/P Rajadram, Senior Lecturer, Research Unit, Department of Surgery, UMMC, Malaysia
4. Professor Dr April Roslani, Head of Department of Surgery, UMMC, Malaysia
5. Professor Dr. Tunku Kamarul Zaman Bin Tunku Zainol Abidin, Director of UMMC, Malaysia

## Reference

1. WHO. Rolling updates on coronavirus disease (COVID-19). WHO 2020.

2. Wu, Z. and J.M. McGoogan, Characteristics of and Important Lessons From the Coronavirus Disease 2019 (COVID-19) Outbreak in China: Summary of a Report of 72314 Cases From the Chinese Center for Disease Control and Prevention. JAMA, 2020.

3. Emanuel, E.J., et al., Fair Allocation of Scarce Medical Resources in the Time of Covid-19. N Engl J Med, 2020. 382(21): p. 2049–2055.

4. Tang, K.H.D., Movement control as an effective measure against Covid-19 spread in Malaysia: an overview. Journal of Public Health, 2020.

5. Kaye, K., et al., Elective, Non-urgent Procedures and Aesthetic Surgery in the Wake of SARS-COVID-19: Considerations Regarding Safety, Feasibility and Impact on Clinical Management. Aesthetic Plast Surg, 2020.

6. Tay, K., et al., COVID-19 in Singapore and Malaysia: Rising to the Challenges of Orthopaedic Practice in an Evolving Pandemic. Malays Orthop J, 2020. 14(2).

7. Al-Jabir, A., et al., Impact of the Coronavirus (COVID-19) pandemic on surgical practice - Part 1 (Review Article). Int J Surg, 2020.

8. Pather, N., et al., Forced Disruption of Anatomy Education in Australia and New Zealand: An Acute Response to the Covid-19 Pandemic. Anat Sci Educ, 2020. 13(3): p. 284–300.

9. Li, L., Q. Xv, and J. Yan, COVID-19: the need for continuous medical education and training. Lancet Respir Med, 2020. 8(4): p. e23.

10. Liang, Z.C., S.B.S. Ooi, and W. Wang, Pandemics and Their Impact on Medical Training: Lessons From Singapore. Acad Med, 2020.

11. Dedeilia, A., et al., Medical and Surgical Education Challenges and Innovations in the COVID-19 Era: A Systematic Review. In Vivo, 2020. 34(3 Suppl): p. 1603–1611.

12. Ministry of Health, M., Housemanship. 2016.

13. Gopalakrishnan, U., Affan, Zamri, Kamal, Sandheep Stress perceived by houseman in a hospital in northern Malaysia. The Medical Journal of Malaysia, 2016. 71(1):8-11(71(1):8-11).

14. Al-Dubai, S.A., et al., Emotional burnout, perceived sources of job stress, professional fulfillment, and engagement among medical residents in Malaysia. ScientificWorldJournal, 2013. 2013: p. 137620.

15. Ministry of Health, M., Covid-19: 1,000 housemen to be stationed nationwide to support medical staff. The Star, Saturday, 14 Mar 2020.

16. Tan, B.Y.Q., et al., Psychological Impact of the COVID-19 Pandemic on Health Care Workers in Singapore. Ann Intern Med, 2020.

17. Zhang, W.R., et al., Mental Health and Psychosocial Problems of Medical Health Workers during the COVID-19 Epidemic in China. Psychother Psychosom, 2020: p. 1–9.

18. Wong, J.E.L., Y.S. Leo, and C.C. Tan, COVID-19 in Singapore-Current Experience: Critical Global Issues That Require Attention and Action. JAMA, 2020.

19. Kogan, M., et al., Orthopaedic Education During the COVID-19 Pandemic. J Am Acad Orthop Surg, 2020. 28(11): p. e456–e464.

20. Gallagher, T.H. and A.M. Schleyer, “We Signed Up for This!” - Student and Trainee Responses to the Covid-19 Pandemic. N Engl J Med, 2020.

21. Maunula, L., The pandemic subject: Canadian pandemic plans and communicating with the public about an influenza pandemic. Healthc Policy, 2013. 9(Spec Issue): p. 14–25.

22. Ranney, M.L., V. Griffeth, and A.K. Jha, Critical Supply Shortages - The Need for Ventilators and Personal Protective Equipment during the Covid-19 Pandemic. N Engl J Med, 2020. 382(18): p. e41.

23. Ahmed, H., M. Allaf, and H. Elghazaly, COVID-19 and medical education. Lancet Infect Dis, 2020. 20(7): p. 777–778.

24. Newman, N.A. and O.M. Lattouf, Coalition for medical education-A call to action: A proposition to adapt clinical medical education to meet the needs of students and other healthcare learners during COVID-19. J Card Surg, 2020. 35(6): p. 1174–1175.

25. Aiello, A., et al., Resilience training for hospital workers in anticipation of an influenza pandemic. J Contin Educ Health Prof, 2011. 31(1): p. 15–20.

